# Open-source analytics tools for studying the COVID-19 coronavirus outbreak

**DOI:** 10.1101/2020.02.25.20027433

**Authors:** Tianzhi Wu, Erqiang Hu, Xijin Ge, Guangchuang Yu

## Abstract

The COVID-19 outbreak originated at the end of 2019 from Wuhan [1, 2], a city in Hubei province in central China. According to the World Health Organization (WHO), there were 88,948 confirmed cases and 3,043 deaths from 65 countries as of March 2, 2020. In China, the outbreak has effectively confined over 1 billion people to their apartments and homes since the end of January 2020 and continues to disrupt healthcare, wellbeing, and the economy. As the situation in China appears to be stabilizing, sharp increases in confirmed cases are being reported in South Korea, Italy, Japan, and Iran.

---

Access to real-time data is essential to the public, scientists, and public health officials. The interactive dashboard developed by Dong *et al*. [3] is a powerful tool to assess the current situation. To provide direct access to real-time epidemiological data on this outbreak, we developed an R package, nCov2019 [4]. This open-source software aggregates data from four different sources. We retrieve the current numbers of confirmed cases and deaths in geographical locations using API (application programming interface) calls to the Tencent SARS-COV-2 website [5]. Updated several times a day, the Tencent website relies on official data obtained from the Chinese provincial health agencies, China National Health Commission (CNHC), the World Health Organization (WHO), and public health agencies in other countries. More importantly, our R package offers access to three data sources with detailed daily statistics from December 1, 2019, for 43 countries and more than 500 Chinese cities. Our first source is obtained directly from CNHC, which is official historical statistics for the 34 the Chinese provinces and special districts. The second source is from a non-governmental organization Dingxiangyuan [6], which has been continuously aggregating official data from provincial and city health agencies and the CNHC. The third source is a public GitHub repository [7], which derives data from the literature [8] for December 1, 2019, to January 10, 2020, after which it relies on the Chinese news aggregator Toutiao API. This GitHub repository includes historical data for Chinese cities as well as 43 countries. All datasets are updated daily, and they are consistent with each other (see Suppl. Doc. 1).

As demonstrated in Suppl. Doc. 1, this new package also contains functionalities to facilitate data visualization. For example, with one command, users can easily plot the distribution of cases on the maps of the world, China, and even individual provinces (Figure 1). With historical data, we can incorporate temporal and spatial information to create an animation to help us understand disease transmission and examine the spread of the COVID-19 outbreak.

**Figure 1.**
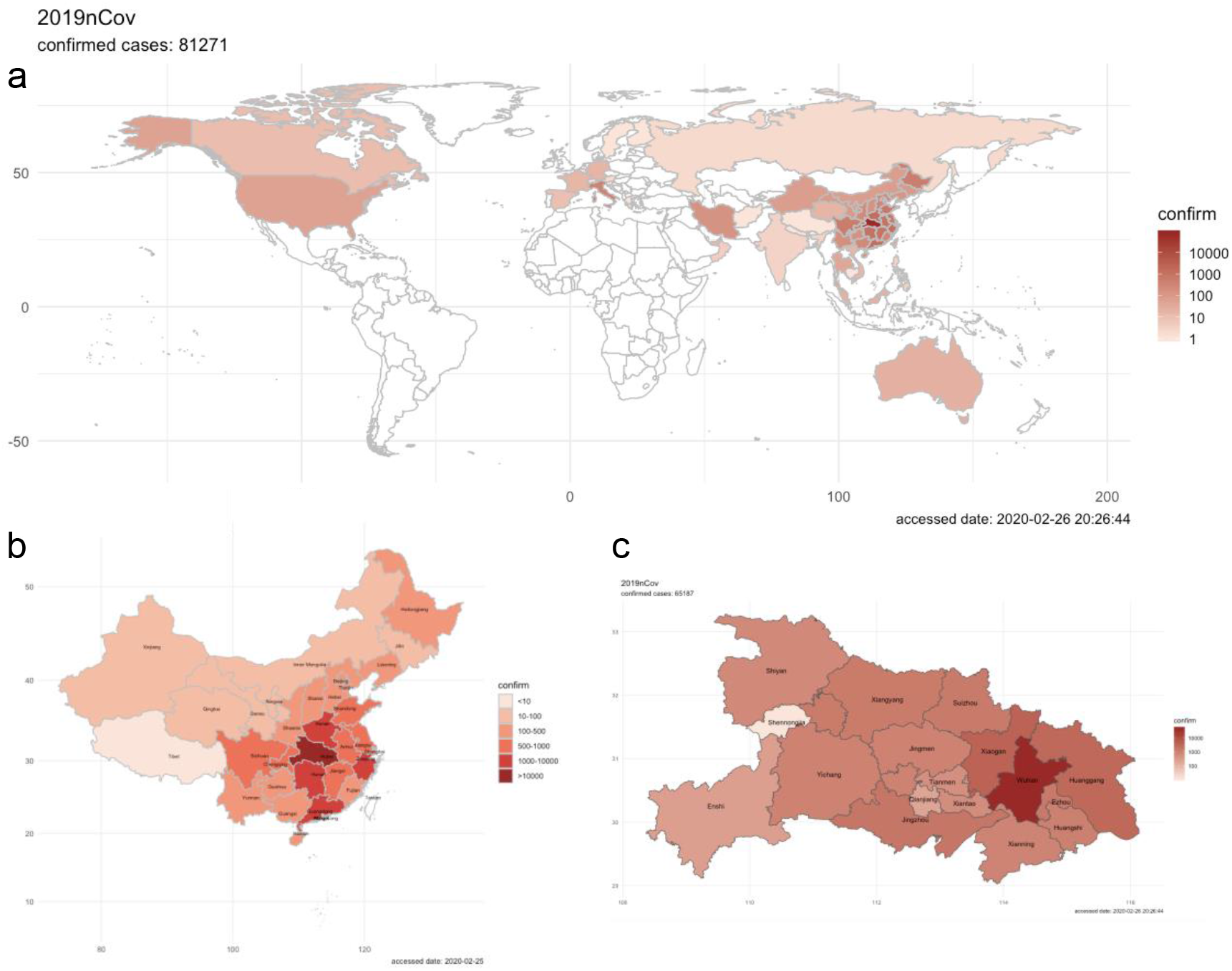
a) Number of confirmed COVID-19 cases across different countries (a), Chinese provinces (b), and cities (c).

To enable users to access these datasets without coding, we also developed interactive web apps in both English [9] and Chinese [10]. As demonstrated in Supp. Doc. 1, these apps can also be run locally from Rstudio. Using these apps, users can gain insights by quickly generating all 23 plots in Supp. Doc. 2 based on daily updated data. Complementing the dashboard by Dong *et al*. [3], our web app enables users to select their regions of interest and check both the historical and real-time data. Generated by the app on February 25, 2020, Figure 2 shows that the total confirmed cases in the provinces outside Hubei are stabilizing, following a similar trend. The extreme measures that the Chinese government took since January 23 seem to be working.

**Figure 2.**
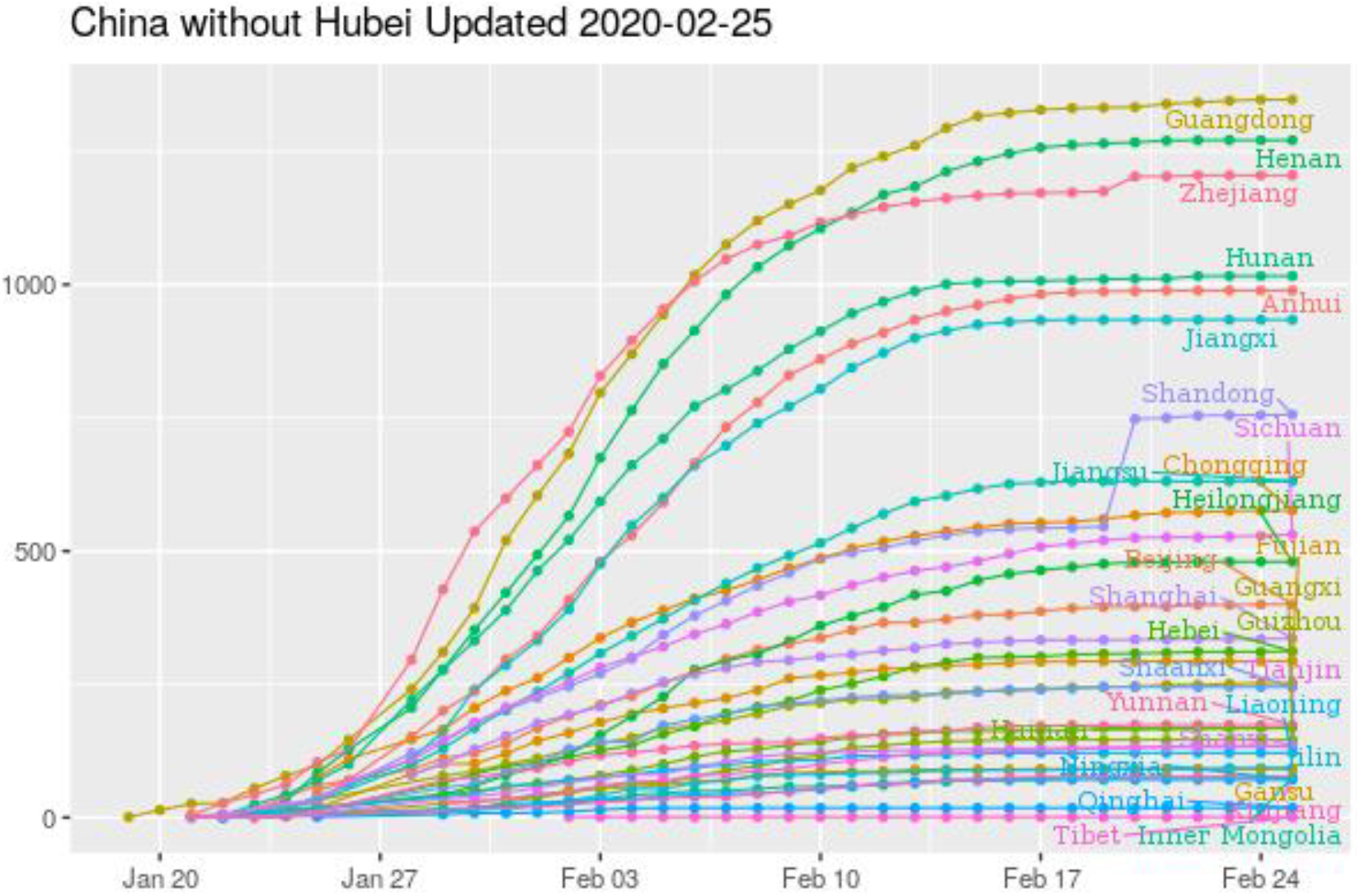
The number of confirmed COVID-19 cases in Chinese provinces except for Hubei. Through the interactive website, users can get detailed statistics for all provinces and cities in China.

Built with the RStudio Shiny framework, these apps contain a simple forecast module. We first converted the log-transformed numbers of cases or deaths as a time-series data, then used the exponential smoothing method (*ets*) in the R package ***forecast*** [11] with default settings to forecast the total cases. On February 7, 2020, this simple model predicted that the death toll would reach 2000 in ten days, a staggering number at the time that later materialized, unfortunately. We also converted the raw number of cases as percent daily changes and conducted a similar forecast. Interestingly, daily percent changes in both confirmed cases and deaths in China are decreasing linearly except for a few outliers (see Figure 16 and 18 in Supplementary Document 2).

Even though not all data sources are official statistics, this kind of detailed data offers a unique opportunity to study this novel pathogen. The hundreds of cities could even be considered as semi-independent outbreaks, as many of them are far from the epicenter and effectively on lockdown from the end of January 2020. As shown in Figures 5 and 6 in Supp. Doc. 2, the death rate, estimated by dividing current total deaths by total confirmed cases, in Wuhan is 4.47%. Probably due to an overwhelmed healthcare system, this death rate is higher than the average of 2.92% (95% confidence interval [2.35% - 3.38%]) observed in 22 Chinese cities with 200 or more confirmed cases. Cities in Hubei province have higher fatality rates than cities in other regions (Figure 6 in Supp. Doc. 2). Internationally, the death rate in Japan (2.50%) is close to that of Italy (2.60%), lower than the 3.67% observed in China overall (Figure 17 in Supp. Doc. 2). The death rate in Iran is 9.63%, probably due to underreported cases.

The rapid, exponential growth phase in China spans roughly from January 15 to February 15, 2020, when the number of confirmed cases skyrocketed 1670-fold from 41 to 68,500. Such rapid growth is now evident in South Korea, Italy, and Iran (Figure 3). Other countries with a smaller number of cases but showing a sharp upward trend include Germany, Spain, and France. If not managed well, tens of thousands of cases in each of these and other countries could be possible in weeks. Public health officials need to grasp the power of exponential growth.

**Figure 3.**
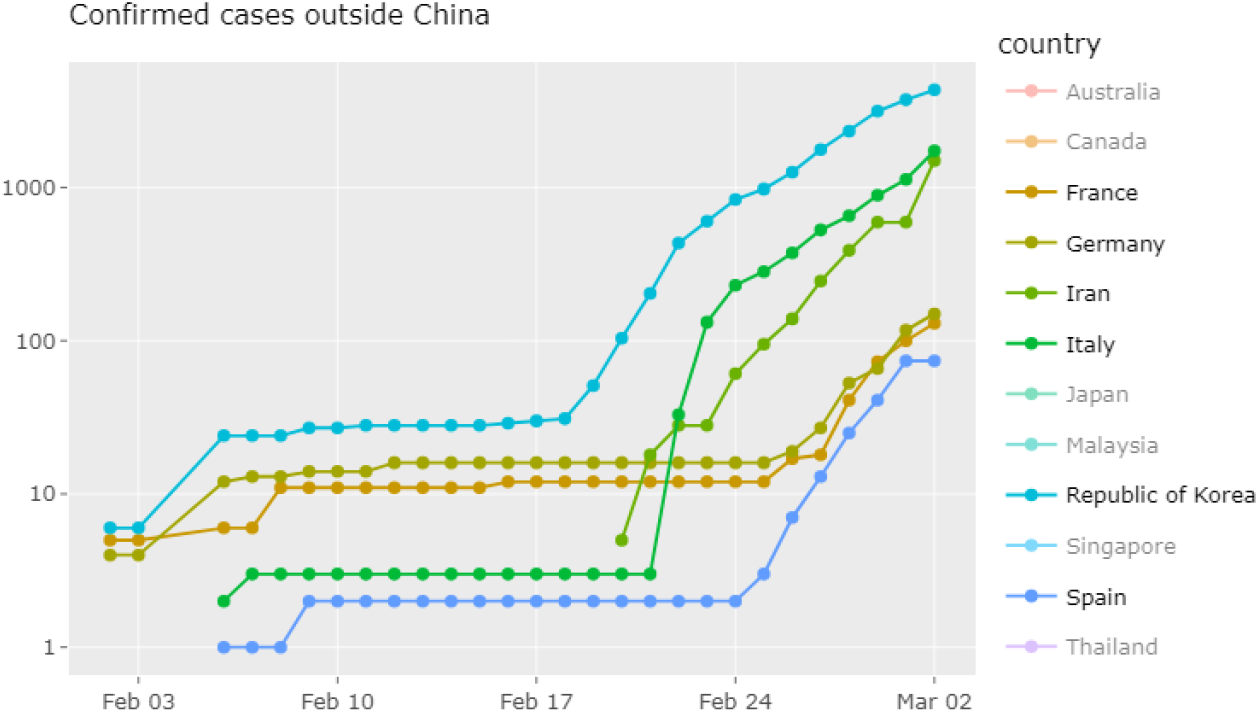
Countries with rapidly growing COVID-19 cases. This plot is obtained using our interactive app.

Currently, city-level historical data is only available for China. These data sources occasionally change data formats, which requires us to monitoring the data sources. If the APIs stopped providing data, the real-time data would not be updated. But the historical data will remain accessible for researchers. We will maintain the web apps during this outbreak.

Our nCov2019 package reduces the barrier for researchers and public health officials in obtaining comprehensive, up-to-date data on this ongoing outbreak. With this package, epidemiologists and other scientists can directly access data from four sources, facilitating mathematical modeling and forecasting of the COVID-19 outbreak. The interactive web apps are accessible to the general public and could also be easily customized by researchers to produce other dashboards or track other countries. We hope these analytics tools could be useful in studying and managing this pathogen on a global scale.

## Data Availability

Source code and data are publically available on GitHub.

https://github.com/GuangchuangYu/nCov2019

Supplementary Document 1: Detailed tutorial and example of how to use the R package.

Supplementary Document 2: Example of plots obtained from our web app.

